# A comprehensive digital phenotype for postpartum hemorrhage

**DOI:** 10.1101/2021.03.01.21252691

**Authors:** Amanda B Zheutlin, Luciana Vieira, Shilong Li, Zichen Wang, Emilio Schadt, Susan Gross, Joanne Stone, Eric Schadt, Li Li

**Affiliations:** Clinical Informatics, Sema4, 333 Ludlow St., Stamford, CT 06902, USA; Division of Maternal Fetal Medicine, Department of Obstetrics, Gynecology, and Reproductive Science, Icahn School of Medicine at Mount Sinai, New York, NY 10029, USA; Department of Genetics and Genomic Sciences, Icahn School of Medicine at Mount Sinai, New York, NY 10029, USA

**Author notes:** Equal contribution (Amanda B Zheutlin, Luciana Vieira, Eric Schadt, Li Li). Corresponding Author: Li Li, M.D., 333 Ludlow St., North Tower, 7^th^ Floor, Stamford, CT 06902.

**Keywords:** postpartum hemorrhage, digital phenotype, electronic medical records, maternal morbidity

## Abstract

**Objective:** We aimed to establish a comprehensive digital phenotype for postpartum hemorrhage (PPH). Current guidelines rely primarily on estimates of blood loss, which can be inaccurate and biased, and ignore a suite of complementary information readily available in electronic medical records (EMR). Inaccurate and incomplete phenotyping contribute to ongoing challenges to track PPH outcomes, develop more accurate risk assessments, and identify novel interventions.

**Methods:** We constructed a cohort of 71,944 deliveries from the Mount Sinai Health System, 2011-2019. Estimates of postpartum blood loss, shifts in hematocrit intra- and postpartum, administration of uterotonics, surgical treatments, and associated diagnostic codes were combined to identify PPH retrospectively. All clinical features were extracted from structured EMR data and mapped to common data models for maximum interoperability across hospitals. Blinded chart review was done on a randomly selected subset of cases and controls for validation and performance was compared to alternate PPH phenotypes.

**Results:** We identified 6,639 cases (9% prevalence) using our phenotype – more than three times as many as using blood loss alone (N=1,747), supporting the need to incorporate other diagnostic and treatment data. Blinded chart review revealed our phenotype had 96% sensitivity, 89% precision, 77% specificity, and 89% accuracy to detect PPH. Alternate phenotypes were less accurate, including a common blood loss-based definition (67%) and a previously published digital phenotype (74%).

**Conclusion:** We have developed a scalable, accurate, and valid digital phenotype that may be of significant use for tracking outcomes and ongoing clinical research to deliver better preventative interventions for PPH.

## BACKGROUND AND SIGNIFICANCE

Postpartum hemorrhage (PPH) is a leading cause of maternal mortality in the US[1,2]. The majority of these deaths are preventable and a primary cause is error or delay in diagnosis and treatment[2–6]. Though mortality rates due to PPH have remained stable over the past 15 years[7,8], the prevalence has increased considerably[9], as well as the need for critical interventions to treat severe cases[1,9–12]. Even in developed countries where rates are significantly lower than in developing countries[13], prevalence estimates range from 3% to 9% depending on the definition[7,11,14–17]. There is a critical need for optimization of preventative care (e.g., earlier identification of risk factors and prompt recognition of PPH) and treatment modalities to reduce morbidity and mortality[18].

Historically, postpartum hemorrhage has lacked a single, consistent definition and has relied heavily on visual estimates of blood loss, which can be inaccurate, biased, and unreliable[6,18–23]. More recently, in an effort to standardize clinical obstetric definitions, the American College of Obstetricians and Gynecologists (ACOG) developed the reVITALize program, which defines postpartum hemorrhage as a cumulative blood loss of greater than or equal 1000mL or blood loss accompanied by signs or symptoms of hypovolemia within 24 hours following delivery (including intrapartum blood loss)[24]. However, blood loss is still most commonly estimated visually and this measure is central to diagnosis and initiation of treatment[4,6], as well as the primary metric used retrospectively for hospital quality outcomes and research aimed at improving prevention and treatment of PPH[15]. Overreliance on estimated blood loss (EBL) alone has contributed to underestimation of hemorrhage[17,19,20,22,23,25]; this is a key area for improvement in PPH prevention efforts.

Due to these limitations, practice is shifting towards more objective measurements, such as quantitative blood loss (QBL), with obstetric hemorrhage toolkits like the California Maternal Quality Care Collaborative[26]. The use of QBL can lead to earlier interventions and a greater emphasis on attention to blood loss, though these practices have not been widely adopted since they require hospitals to have specialized equipment and training for providers[6,21]. Additionally, while quantitative measures are more accurate than visual estimates, they can still be imprecise, as well as variable across caregivers[15,27]. Furthermore, this threshold is necessarily arbitrary and some women needing care for hemorrhage may ultimately lose less than 1000mL[23].

To mitigate limitations of using blood loss alone to definite PPH, additional measures have been proposed to refine case definitions. A proxy measure of blood loss is change in hematocrit values, with a 10% drop indicating PPH[1], although this may have low specificity and is affected by global changes in fluids like dehydration or any infusions[21]. Alternatively, PPH can also be indexed with diagnostic codes or with indications of severe outcomes like blood transfusions or surgical interventions[15,28]. However, diagnostic codes have low sensitivity[16], while indications of severe outcomes have high sensitivity, but occur in a minority of cases. One group has suggested combining multiple retrospective diagnostic indicators with medications to manage uterine atony to improve detection of PPH[15,21]. Uterotonics, including oxytocin, carboprost tromethamine, misoprostol, and methylergonovine, are the first line intervention for acute medical management of PPH[1], so they may be useful markers for identifying PPH. However, given the novelty of the digital phenotype this group detailed, validation against gold standard labels is a crucial step towards widespread adoption.

Here, we aimed to establish a physician-validated, comprehensive digital phenotype for PPH using information extracted from electronic medical records (EMR) in a large US health system. We used several sources to identify deliveries with significant blood loss, as well as deliveries where medical or surgical interventions for treating PPH were given. Through careful extraction of medication dosing and timing, mapping of fluctuations in lab values during labor and delivery, and synthesis of medical observations across labor and delivery admission, we aimed to develop a comprehensive phenotype to retrospectively identify deliveries with PPH. To validate this phenotype, blinded chart review by a physician was conducted on a subset of patients. Performance of our phenotype was then compared to both a common definition of PPH (>= 1000mL blood loss)[1] and the most comprehensive previously developed digital phenotype[15]. Inaccurate phenotyping remains a significant barrier for tracking incidence and management of PPH in hospitals, developing more accurate risk stratification tools, and identifying novel interventions. Our goal was to provide a robust digital phenotype that can be readily implemented retrospectively for both quality improvement initiatives and clinical research.

## METHODS

### Patient population

We used deidentified EMR data provided by the Mount Sinai Data Warehouse (MSDW) from the Mount Sinai Health System (MSHS), one of the largest and most comprehensive EMR systems in New York City. MSHS includes 5 member hospitals with EMR from 2000-2020, which draw from a racially and ethnically diverse patient population. Clinical variables including patient demographics, medical histories, or visit details were available for 9 million unique patients. These deidentified data were used to construct a delivery cohort and develop a digital phenotyping algorithm. A diagram of our workflow is presented in Figure 1A. We received approval from the Icahn School of Medicine at Mount Sinai Institutional Review Board (IRB-17-01245) to conduct this study.

**Figure 1.**
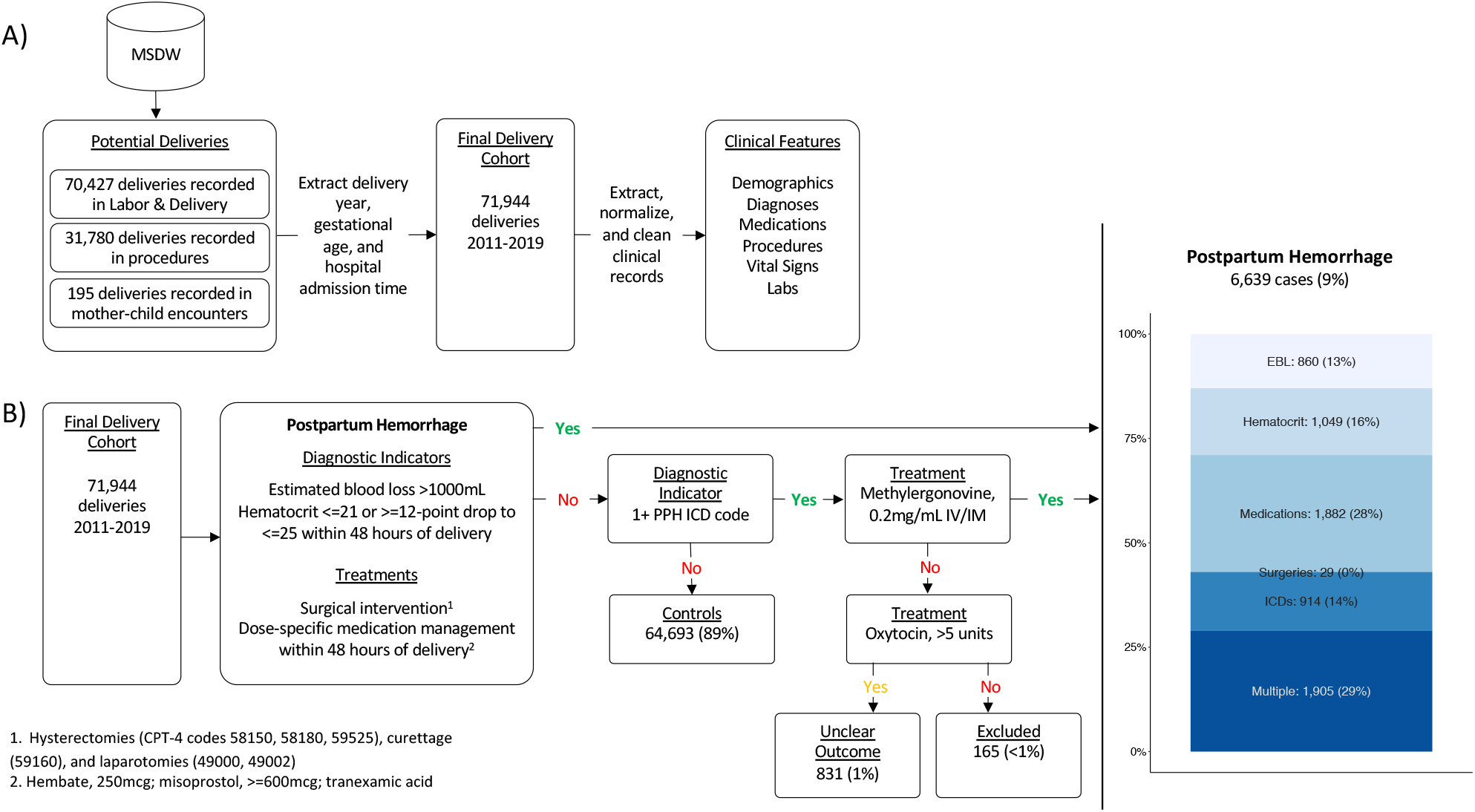
Workflow for data extraction (A) and digital phenotype (B).

### Delivery cohort

To identify all deliveries, we used three sources: 1) a standardized delivery summary completed by delivery staff on Labor & Delivery, 2) procedure records for vaginal or Cesarean deliveries (identified using CPT-4 and ICD-10-PCS billing codes), and 3) linked mother-infant hospital visit records timestamped to the infant’s day of birth. For all deliveries, we identified gestational weeks at delivery, delivery time, delivery method, parity, and hospital admission time. When gestational weeks at delivery was not recorded, it was estimated using gestational age reported for prenatal visits (admit reason). When delivery time was not available, we used the final procedure timestamp or the timestamp at which the mother received 5 or more units of oxytocin (a prophylactic dose given immediately after delivery of the anterior shoulder)[14]. Delivery method was labeled using delivery procedure records and ICD-9-CM or ICD-10-CM diagnostic codes for delivery (70 codes) given to either the mother or infant. Parity was estimated by assuming the first delivery for each woman was the earliest one included in our cohort and that all following deliveries were also at MSHS. Finally, hospital inpatient admission and unit transfer times were extracted to create admission-delivery journeys. Deliveries without any gestational age information or without admission time were excluded. We also limited the cohort to deliveries from January 1, 2011 through December 31, 2019 to ensure records were complete (prior to 2011, data availability through EMR is limited).

### Clinical feature cleaning and normalization

All available demographic information, lab tests, vital signs, diagnoses, medications, and procedures were extracted for all women in our delivery cohort. We standardized these data by mapping native coding systems to common frameworks that are part of the Unified Medical Language System (UMLS) – a process that also increases interoperability between healthcare systems. All available observations were cleaned and normalized within data type.

For patient demographics, we extracted mother’s age at delivery, race, ethnicity, and insurance. When there were inconsistencies within a patient’s history of self-reported race or ethnicity, we assigned the most common self-report. Lab test names and units were mapped to logical observation identifiers names and codes (LOINC). Values were cleaned (invalid results and text removed) and converted to numeric values or standardized to non-numeric scales as appropriate. Duplicate results (e.g., ‘preliminary’ and ‘final’ results from the same test with the same values) were filtered to retain the earliest result. Vital signs, including weight, height, temperature, respirations, pulse, diastolic blood pressure (DBP), and systolic blood pressure (SBP) were standardized to common names and unit scales. Diagnoses from ICD-9-CM and ICD-10-CM were combined via mapping to broader categories using the Clinical Classifications Software. We filtered medications to those administered (i.e., directly given to patients) and mapped all medication names to RxNorm ingredients. Procedures were recorded through CompuRecord, an anesthesia information management program, using CPT-4 codes. When procedures included multiple timepoints (e.g., procedure start, anesthesia given, fluid given), only the earliest one was retained.

#### Clinical and obstetric characteristics

The latest hematocrit (LOINC 20570-8) test result and vital signs given within 48 hours prior to delivery were used as baseline measures. Oxytocin or misoprostol administered after hospital admission and prior to delivery was considered evidence of labor induction or augmentation. 65 ICD-9/ICD-10 codes given during pregnancy or within 30 days post-delivery were used to index pregnancies with multiple gestation.

### Digital phenotyping algorithm for PPH

We aimed to identify women who had significant blood loss, as well as those who had PPH-specific interventions. A diagram of our workflow is presented in Figure 1B.

#### Diagnostic indicators of PPH

We used multiple sources to detect substantial blood loss postpartum. First, we considered EBL or QBL by clinicians post-delivery to have exceeded 1000mL as evidence of PPH. MSHS adopted quantification practices in 2017; blood loss values were estimated prior to then. Since EBL is biased towards underreporting PPH[6,19,20,23], and quantified blood loss is not always reliable[15,27], we also included women with critically low hematocrit (<=21) or a greater than 12-point drop from baseline – a proxy measure for blood loss[1] – that resulted in a minimum value at or below 25 within 48 hours of delivery. Finally, we included women given one or more of 30 diagnostic codes selected by a Maternal Fetal Medicine specialist as indicators of PPH. Since ICD codes can be assigned to a visit after care was given, we considered codes given on delivery day or within the following 14 days to reflect events during delivery. ICD diagnoses are often inaccurate when used on their own[29], so we additionally required administration of any uterotonic medication except oxytocin (carboprost tromethamine, misoprostol, methylergonovine). Oxytocin was excluded due to its routine use in the active management of the third stage of labor[1].

#### Interventions to prevent or treat PPH

We also identified deliveries where uterotonics or surgical interventions intra- or postpartum were given to prevent or treat maternal hemorrhage. Because methylergonovine can be given prophylactically or as treatment, we included women given methylergonovine intramuscularly only if they were also given an ICD code for PPH (as described above). If one first line uterotonic does not sufficiently control bleeding, acute medical management of postpartum hemorrhage requires combination use of misoprostol, carboprost tromethamine, and/or tranexamic acid (an antifibrinolytic drug that promotes blood clotting)[1]. As such, we included women administered 250mcg of carboprost tromethamine, >=600mcg of misoprostol, or any dose of tranexamic acid within 48 hours of delivery[1,14].

When hemorrhage continues despite medical management, Bakri balloon placement and surgical interventions may be required. This can include procedures occurring during laparotomies (CPT-4 codes 49000, 49002) like placement of compression sutures and uterine artery ligation or embolization, curettage (59160), or, typically as a last result, hysterectomies (58150, 58180, 59525)[1,14,30]. We included all women that underwent any of these procedures postpartum.

#### Deliveries with unclear outcomes and controls

There were 831 deliveries given one or more of 30 PPH ICD codes with a postpartum dose of oxytocin (10 or more units) and had no other indication of PPH. Since this is a plausible scenario for PPH and we wanted to maximize our detection of true cases, we selected some of these charts for review, but labeled them as ‘deliveries with unclear outcomes’ (DUO). Deliveries with an ICD code, but no other indication were excluded (N = 165); all remaining deliveries neither classified as a case nor a DUO were classified as controls.

### Descriptive statistics

We used univariate logistic regression to assess statistical differences between cases and controls in age and gestational weeks at delivery, parity, baseline labs and vital signs, and hours from admission to delivery. Proportional differences in race, ethnicity, insurance, delivery method, labor induction or augmentation, and multiple gestation by case status were assessed using chisquare tests for independence. EBL/QBL differences were assessed separately for vaginal and Cesarean deliveries. We used a Bonferroni correction to conservatively control for multiple comparisons. With this adjustment, the significance threshold was set to alpha < 0.001, two-tailed. All statistical analyses were done using the stats package in R (version 3.6.0).

### Chart review

To assess the validity of our digital phenotype, we selected 45 charts labeled using our phenotype to be manually reviewed by a physician who was blind to the phenotype labels. Charts labeled as controls were selected at random and charts labeled as cases were selected randomly within categories to ensure representation of each case indication. In total, there were 12 control charts, 6 DUO charts, and 27 case charts. Within case charts, 8 were identified with EBL, 4 with medications, 6 with drops in hematocrit, 3 with ICDs and methylergonovine, and 6 with more than one indication.

We had two aims for this review. The first was to verify that our data mining techniques were accurate. Our data was deidentified to ensure patient privacy, so it did not include all information recorded for a delivery, including any information from physician or surgical notes. Thus, we wanted to verify data accuracy by comparing information extracted from EMR including lab results, medications, procedures, delivery time and method, and blood loss values to what was available in native clinical charts accessible by physicians. We summarized data accuracy by overall rates of exact matching values between our data and chart review.

Our second aim was to assess the validity of our digital phenotyping algorithm. Charts were reviewed for evidence of significant blood loss as indicated in notes or lab tests, explicit indication of PPH in delivery summary or visit notes, or evidence of interventions specifically for managing PPH clearly beyond standard care. A judgement for each chart of ‘yes’, ‘no’, or ‘unclear’ was made based on this evidence. We summarized the performance of our digital phenotype by calculating specificity, sensitivity, positive and negative precision, and accuracy of our labels compared to chart review labels.

For comparison, we also calculated these same metrics for each criterion individually (i.e., by using presence or absence of that criterion as the case label and comparing it to chart review labels) in order to assess the value of combining criteria relative to each indicator on its own. Finally, to compare our digital phenotype with alternate phenotypes, we generated case labels based on the EBL criterion from the ACOG guidelines[1] (cases defined as any delivery with EBL/QBL >=1000mL) and the most comprehensive previously published digital phenotype, which was not evaluated with chart review[15]. ACOG additionally defines any blood loss followed by signs or symptoms of hypovolemia to be PPH, however, this criterion is difficult to ascertain from EHR data without discrete definitions. Because there is significant variability in the clinical and vital signs changes that are associated with blood loss, there are no established cutoff points to trigger clinical interventions[31,32]. The latter phenotype was originally proposed as four mutually exclusive levels of risk, which we have combined them here into a single phenotype for simplicity. Cases were defined as having any of the following criteria: administration of any uterotonic (except oxytocin), one or more of 12 ICD-9 or ICD-10 codes for PPH, transfusion of blood intra- or postpartum, receipt of intrauterine tamponade device, or hysterectomy. All patient charts were used for comparison performance metrics, regardless of their label using our digital phenotype.

## RESULTS

### Demographic, clinical, and obstetric characteristics for the pregnancy cohort

We identified 73,025 deliveries occurring between January 1, 2011 and December 31, 2019. We excluded 1,081 deliveries in which a hospital admission time could not be identified, leaving 71,944 deliveries from 57,151 mothers in our final cohort. Summary statistics for demographic, clinical, and obstetric characteristics for the entire cohort, as well as for cases and controls, are provided in Table 1.

**Table 1.**
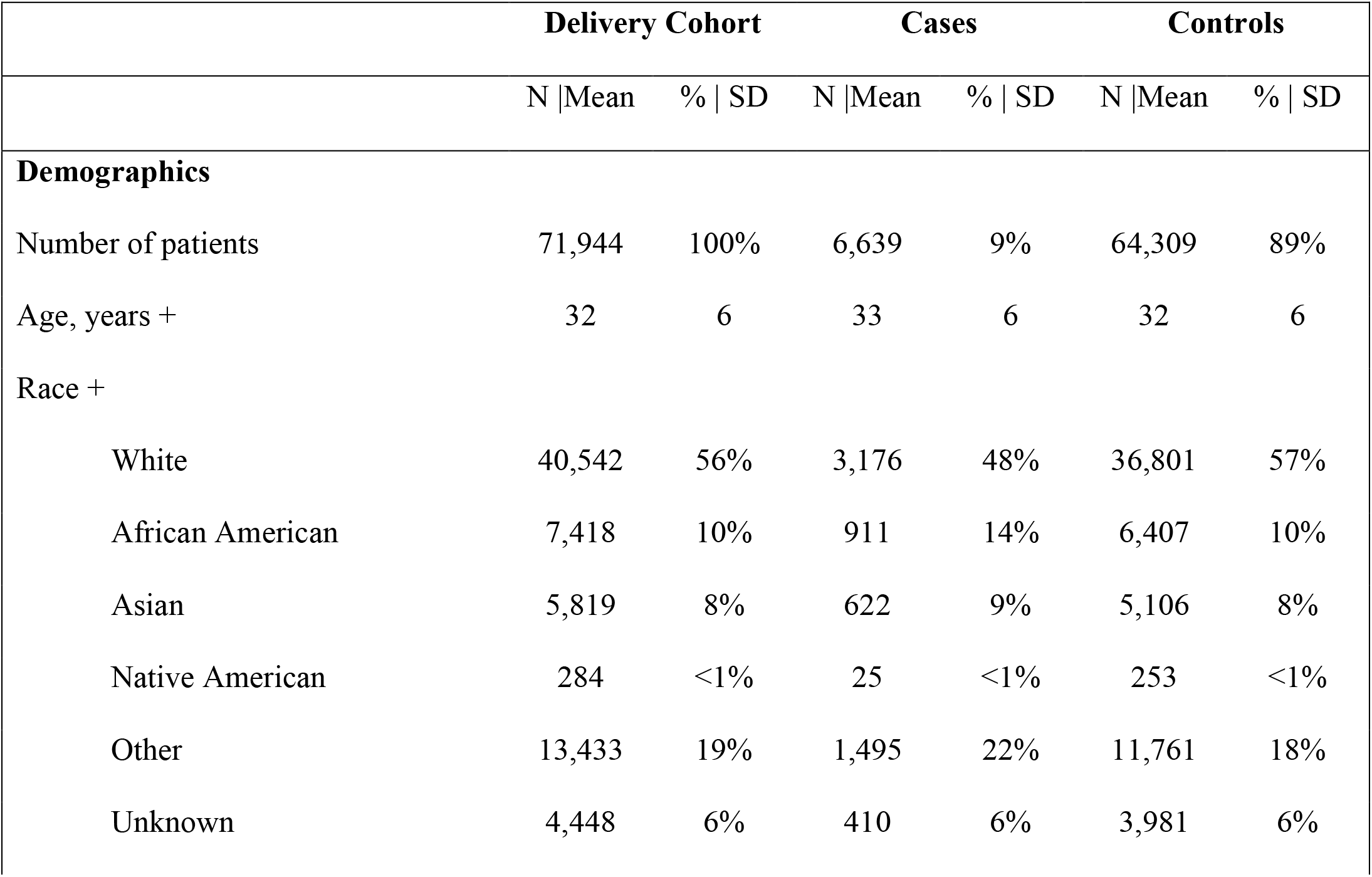
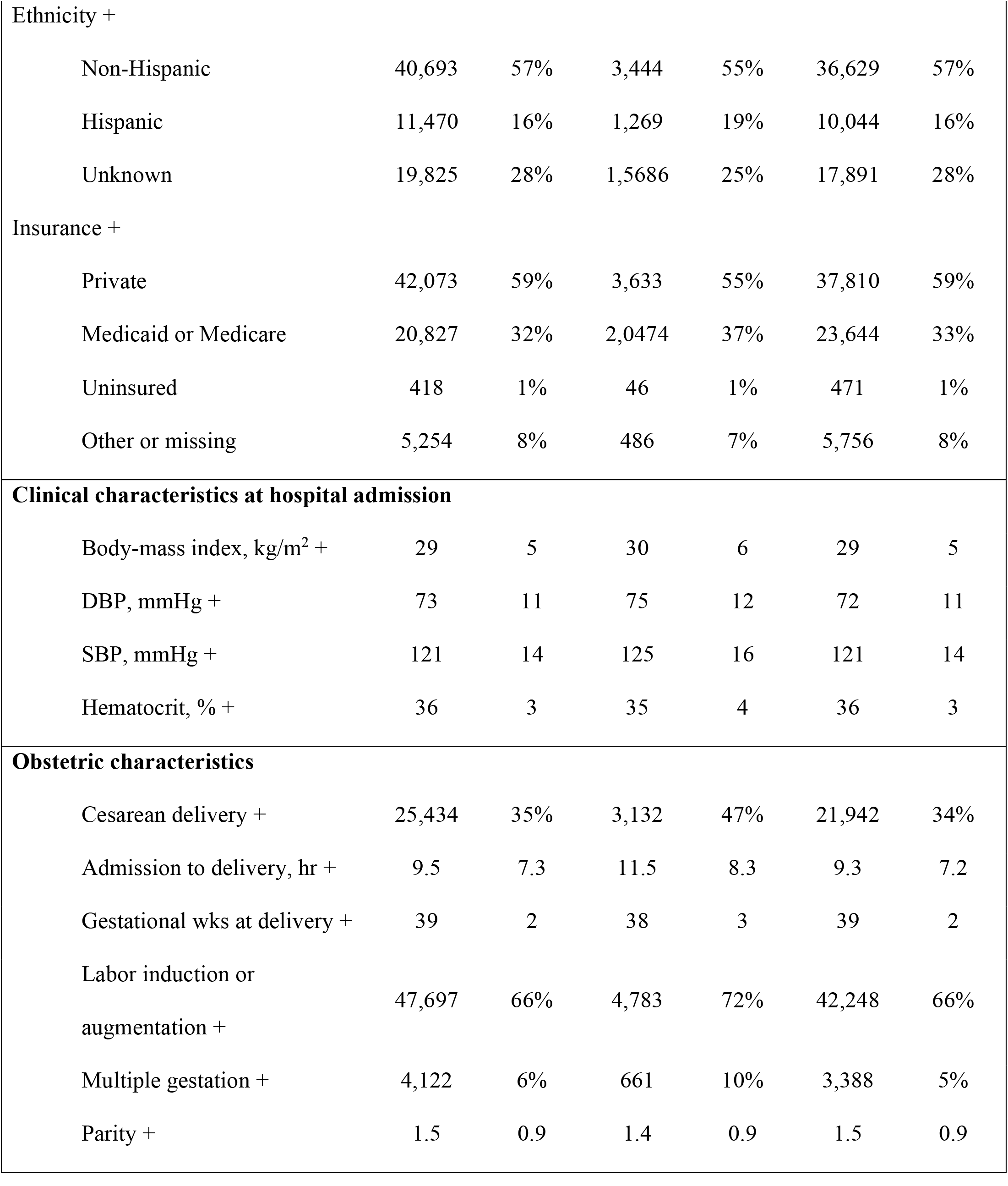
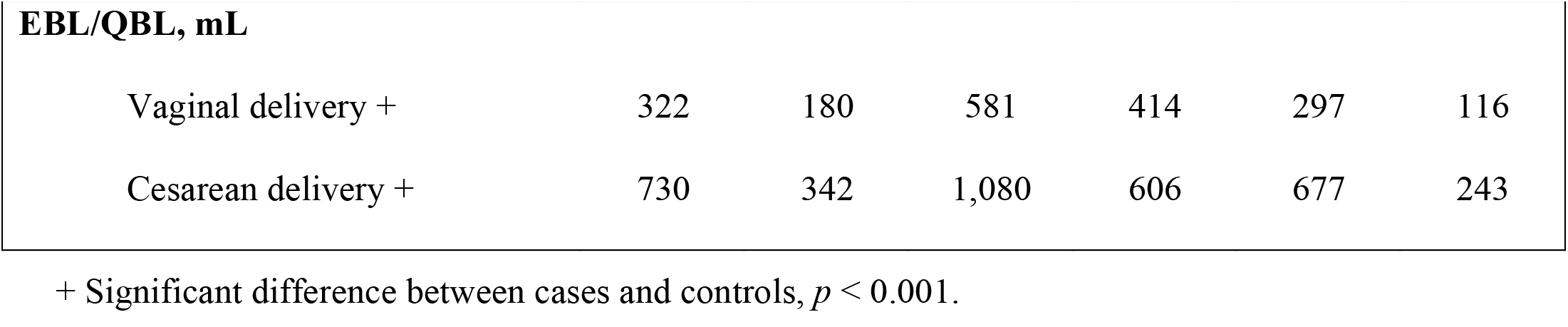
Demographic, clinical, and obstetric characteristics for pregnancy cohort.

### Digital phenotype for PPH

We classified 6,639 deliveries (9% prevalence) as cases using diagnostic indicators of significant blood loss combined with any evidence of PPH-specific interventions. Frequencies for each criterion we considered for inclusion, as well as overall case prevalence are listed in Table 2. We also classified cases based on their indication source (Figure 1B). We found that 71% of cases had evidence from one source of data, including those with multiple indicators within a source (e.g., a woman given carboprost and misoprostol, but no non-pharmaceutical interventions), while 29% of cases had more than one type of indication in their medical record (e.g., at least a 12-point drop in hematocrit to at or below 25 and EBL >1000mL).

**Table 2.** Frequencies of clinical features used to assess medically actionable risk for PPH.

| Criteria | Case* | N | Percent |
| --- | --- | --- | --- |
| EBL/QBL |  |  |  |
| Any record |  | 64,240 | 89% |
| >1000mL | x | 1,747 | 2% |
| Hematocrit |  |  |  |
| Baseline measurement |  | 68,807 | 96% |
| Any follow-up measurement |  | 29,899 | 42% |
| <=21 or >=12-point drop from baseline to<br><=25 | x | 1,949 | 3% |
| Billing codes |  |  |  |
| PPH ICD code |  | 4,219 | 6% |
| PPH ICD code + oxytocin, >=10 units | DUO | 3,829 | 5% |
| PPH ICD code + methylergonovine,<br>0.2mg/mL IM | x | 2,060 | 3% |
| Medication management |  |  |  |
| Oxytocin, >=10 units |  | 61,476 | 85% |
| Methylergonovine, 0.2mg/mL IM |  | 8,094 | 11% |
| Carboprost tromethamine, 250mcg | x | 1,806 | 3% |
| Misoprostol, >=600mcg | x | 2,000 | 3% |
| Tranexamic acid | x | 111 | <1% |
| Surgical interventions |  |  |  |
| Laparotomy | x | 18 | <1% |
| Curettage | x | 51 | <1% |
| Hysterectomy | x | 74 | <1% |
| **Any case indication** | x | **6,639** | **9%** |

\* Indicates case inclusion in digital phenotype (x) or deliveries with unclear outcome (DUO)

### Validation of digital phenotype for PPH

We selected 45 charts to review for data accuracy and digital phenotype validation. Two charts were excluded due to restricted access, resulting in 43 charts for review. Delivery method labels, delivery times, estimates of blood loss, baseline and follow-up hematocrit lab values, and uterotonics administration exactly matched those found in chart review, with the exception of one missing medication for two deliveries. For these cases, methylergonovine was listed only in the patient’s delivery summary, but not in the patient’s medication records, so these events were not found in the structured data extracted from EMR.

We also evaluated the accuracy of our case, control, and DUO labels relative to chart review. Among patients labeled as cases or controls by our digital phenotype (N = 37), 24 had PPH and 13 did not according to physician review. One true PPH case and three controls were misclassified by our phenotype algorithm, yielding 89% accuracy (Table 3). Among patients with unclear outcomes (DUO group), three had PPH and three did not. Overall performance and accuracy by case criterion were detailed in Table 4. We also compared accuracy of our digital phenotype to labels using only one of the criteria we considered (rather than combining them), and two alternate phenotypes: ACOG’s EBL PPH guideline, and the most comprehensive EHR-based digital phenotype previously proposed. All were less accurate than our digital phenotype (Table 3).

**Table 3.** Performance for digital phenotype, individual criteria, and alternate phenotypes

|  | **Sensitivity**<br>**(Recall)** | **Specificity** | **Precision**<br>**(PPV)** | **NPV** | **Accuracy** |
| --- | --- | --- | --- | --- | --- |
| **Digital Phenotype** |  |  |  |  |  |
| Case vs. control | 96% | 77% | 89% | 91% | 89% |
| Case and DUO vs. control | 96% | 63% | 81% | 91% | 84% |
| Case vs. control and DUO | 85% | 81% | 89% | 76% | 84% |

| Phenotype Criterion Alone |  |  |  |  |  |
| --- | --- | --- | --- | --- | --- |
| EBL/QBL >1000mL | 44% | 100% | 100% | 48% | 65% |
| Hematocrit <=21 or 12-point drop to <=25 | 30% | 100% | 100% | 46% | 56% |
| ICD | 74% | 75% | 83% | 63% | 74% |
| Medication (carboprost, misoprostol, tranexamic acid) | 19% | 88% | 71% | 39% | 44% |
| Procedure | 4% | 100% | 100% | 38% | 40% |
| Alternate Phenotype |  |  |  |  |  |
| *Goffman et al.*, levels 1-4 | 85% | 56% | 77% | 69% | 74% |
| ACOG EBL | 59% | 81% | 84% | 54% | 67% |

**Table 4.** Digital phenotype performance by case criterion

| Digital Phenotype |  |  | Physician Label from Chart Review |  |  |
| --- | --- | --- | --- | --- | --- |
| Label | N | Criterion | Case | Control | Unclear |
| **Case** | **26** | **All indications** | **23** | **3** | **0** |
|  | 7 | EBL/QBL | 7 | 0 | 0 |
|  | 3 | ICD + methylergonovine | 2 | 1 | 0 |
|  | 6 | Hematocrit drop or critically low | 6 | 0 | 0 |
|  | 4 | Medication management | 2 | 2 | 0 |
|  | 6 | More than one indication | 6 | 0 | 0 |
| **Control** | **11** | **No case indications** | **1** | **10** | **0** |
| **Case and control** | **37** | **All cases and controls** | **24** | **13** | **0** |
| **DUO** | **6** | **ICD + oxytocin** | **3** | **3** | **0** |

## DISCUSSION

We have developed a comprehensive digital phenotype for PPH using a large and diverse EMR system from New York. Chart review confirmed its validity with 96% sensitivity, 77% specificity, and 89% precision. Considering this performance relative to any individual criterion (accuracy ranging from 40%-74%), the most comprehensive previously proposed digital phenotype (74% accuracy), and a common definition based on the EBL criterion from ACOG’s guidelines (67% accuracy), our digital phenotype affords the highest accuracy (89%).

Concretely, this increase in accuracy allows us to identify more than three times as many cases (N = 6,639) than would be identified using blood loss alone (N = 1,747). While additional clinical criteria may be considered by a clinician when they are determining patient care, for retrospective PPH outcomes assessments and research, EBL is often the objective measure that is consistently documented. Our digital phenotype suggests that this approach may substantially underestimate the incidence.

We also confirmed the data we extracted were highly consistent with clinical notes, highlighting the reliability of our approach. To increase accessibility across healthcare systems, our digital phenotype used only structured data mapped to common data models and did not require the use of advanced methods to mine notes (e.g., natural language processing) or individual chart review to extract data, both of which are variable and time consuming, but are commonly included in phenotyping algorithms[29,33]. Together, we suggest this digital phenotype is a scalable, accurate, and valid research tool that could be used to improve tracking of PPH incidence and management, as well as facilitate research to enhance risk assessment and intervention.

Interestingly, we found that overlap between case-inclusion criteria was only moderate. While 29% of cases had more than one indication (e.g., medication and blood loss >1000mL), most had indications from only one category, underlining the need to incorporate multiple sources of information for identification of PPH cases (Figure 1B). Our most accurate categories on their own were EBL/QBL and hematocrit, which were both 100% accurate (equally accurate as having more than one indication) (Tables 3–4). EBL measures are known to be biased towards underreporting, which is consistent with our findings[6,19,20,23]. Deliveries where blood loss was estimated to be >1000mL were highly likely to be cases, but deliveries with estimated blood loss at or below 1000mL were not always controls (Tables 3–4). One metric for capturing PPH cases with blood loss estimates less than 1000mL may be to use a conservative threshold for hematocrit (12-point drop to <=25). While hematocrit measures can reflect other changes besides blood loss (e.g., administration of intravenous fluids or blood transfusions)[15], we found this threshold to be a good discriminator of cases from controls. Finally, we found that the accuracy of diagnostic billing codes depended on the context. While they were frequently assigned correctly (83% precision; Table 3), precision dropped to 67% with methylergonovine only and to 50% when paired exclusively with oxytocin (Table 4).

This phenotype should be considered in the context of several limitations. We reported three false positives and one false negative in chart review. Of the false positives, two of the three were cases where PPH treatment was applied prophylactically, despite no evidence of hemorrhage, because they had significant risk factors at hospital admission. Considering this, the use of our phenotype may be best suited for identifying women who needed interventions for PPH (using this as our definition, precision would be 96%), although its precision for PPH is still high (89%; Table 3). The false negative was a case with an unanticipated surgical complication, which is a less common cause of PPH (and one not treated with uterotonics), and whose hematocrit values narrowly missed our threshold. While 70-80% of PPH cases are caused by uterine atony, uterine trauma (e.g., lacerations or other tissue tears), retained tissue (e.g., invasive placenta), and acquired or chronic coagulopathies can also cause PPH[1,14]. It is possible that information pertaining to these causes is more readily available through physician notes or blood bank records, which we could not access with deidentified data. However, in general, these causes are less likely to be preventable with improved risk prediction than uterine atony – e.g., intraoperative complications can be unforeseen and thus harder to prevent, whereas atony can be targeted prophylactically with uterotonics – so, again, this suggests the phenotype may be best suited for identifying women likely to benefit from interventions for PPH.

Accurate phenotyping is a cornerstone of high caliber clinical research and hospital quality improvement. Here, we offer a robust, portable, physician-validated digital phenotype for PPH that captures more than three times as many cases as the most commonly used approach by leveraging a suite of complementary information available in EMR. This research tool may be of significant use in designing patient safety initiatives in addition to ongoing clinical research to deliver better preventative interventions for the leading cause of maternal morbidity worldwide.

## Data Availability

Data cannot be shared for ethical/privacy reasons.

## ACKNOWLEDGEMENTS

We would like to thank Mount Sinai Data Warehouse physician team for validating data accuracy and facilitating the chart review process. We would also thank the Sema4 IT team for infrastructural and computational support.

## DATA AVAILABILITY STATEMENT

Data cannot be shared for ethical/privacy reasons.

---

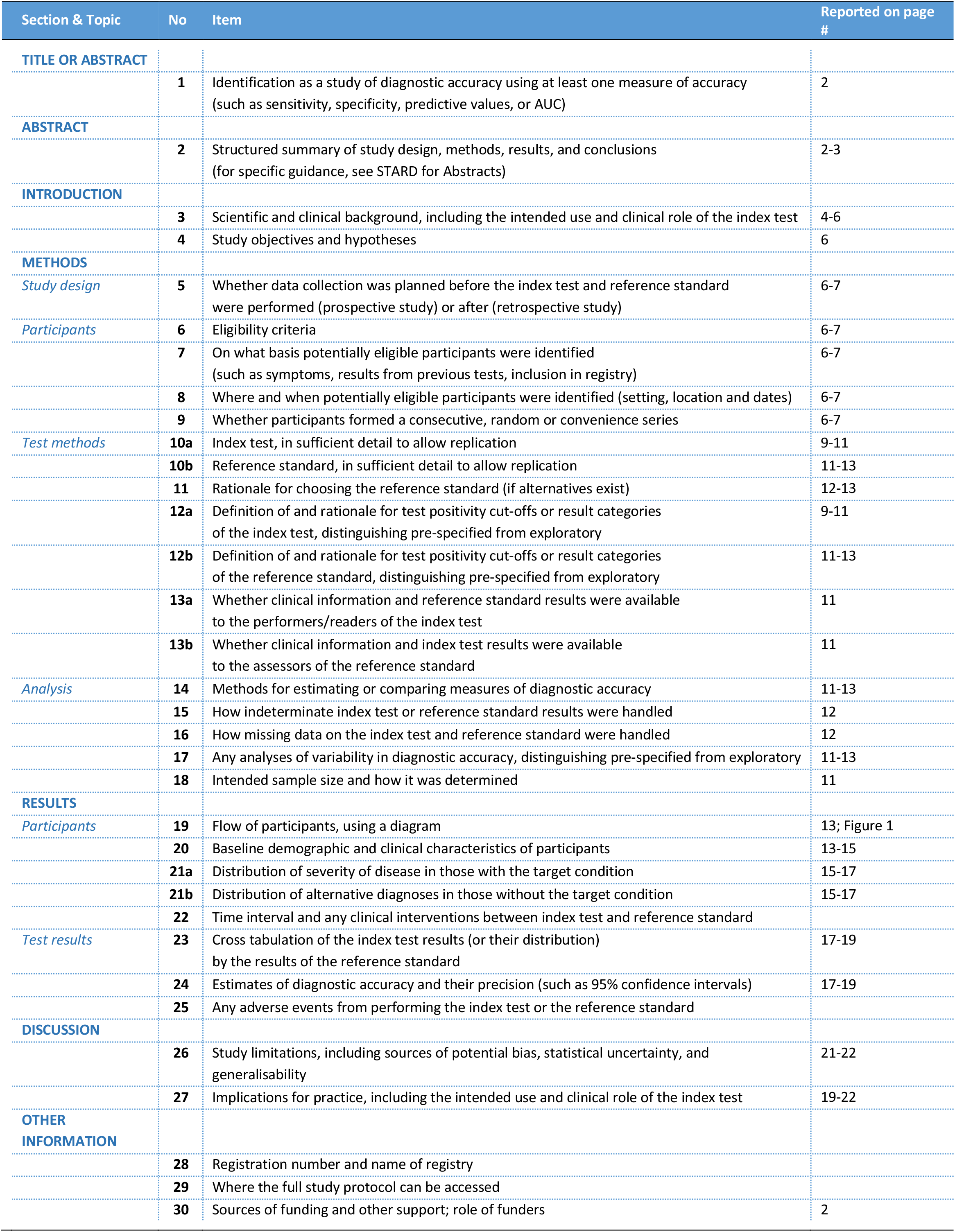

## STARD 2015

### AIM

STARD stands for “Standards for Reporting Diagnostic accuracy studies”. This list of items was developed to contribute to the completeness and transparency of reporting of diagnostic accuracy studies. Authors can use the list to write informative study reports. Editors and peer-reviewers can use it to evaluate whether the information has been included in manuscripts submitted for publication.

### EXPLANATION

A **diagnostic accuracy study** evaluates the ability of one or more medical tests to correctly classify study participants as having a **target condition**. This can be a disease, a disease stage, response or benefit from therapy, or an event or condition in the future. A medical test can be an imaging procedure, a laboratory test, elements from history and physical examination, a combination of these, or any other method for collecting information about the current health status of a patient.

The test whose accuracy is evaluated is called **index test**. A study can evaluate the accuracy of one or more index tests. Evaluating the ability of a medical test to correctly classify patients is typically done by comparing the distribution of the index test results with those of the **reference standard**. The reference standard is the best available method for establishing the presence or absence of the target condition. An accuracy study can rely on one or more reference standards.

If test results are categorized as either positive or negative, the cross tabulation of the index test results against those of the reference standard can be used to estimate the **sensitivity** of the index test (the proportion of participants *with* the target condition who have a positive index test), and its **specificity** (the proportion *without* the target condition who have a negative index test). From this cross tabulation (sometimes referred to as the contingency or “2×2” table), several other accuracy statistics can be estimated, such as the positive and negative **predictive values** of the test. Confidence intervals around estimates of accuracy can then be calculated to quantify the statistical **precision** of the measurements.

If the index test results can take more than two values, categorization of test results as positive or negative requires a **test positivity cut-off**. When multiple such cut-offs can be defined, authors can report a receiver operating characteristic (ROC) curve which graphically represents the combination of sensitivity and specificity for each possible test positivity cut-off. The **area under the ROC curve** informs in a single numerical value about the overall diagnostic accuracy of the index test.

The **intended use** of a medical test can be diagnosis, screening, staging, monitoring, surveillance, prediction or prognosis. The **clinical role** of a test explains its position relative to existing tests in the clinical pathway. A replacement test, for example, replaces an existing test. A triage test is used before an existing test; an add-on test is used after an existing test.

Besides diagnostic accuracy, several other outcomes and statistics may be relevant in the evaluation of medical tests. Medical tests can also be used to classify patients for purposes other than diagnosis, such as staging or prognosis. The STARD list was not explicitly developed for these other outcomes, statistics, and study types, although most STARD items would still apply.

### DEVELOPMENT

This STARD list was released in 2015. The 30 items were identified by an international expert group of methodologists, researchers, and editors. The guiding principle in the development of STARD was to select items that, when reported, would help readers to judge the potential for bias in the study, to appraise the applicability of the study findings and the validity of conclusions and recommendations. The list represents an update of the first version, which was published in 2003.

More information can be found on http://www.equator-network.org/reporting-guidelines/stard.

